# Model Calculations of Aerosol Transmission and Infection Risk of COVID-19 in Indoor Environments

**DOI:** 10.1101/2020.09.22.20199489

**Authors:** J. Lelieveld, F. Helleis, S. Borrmann, Y. Cheng, F. Drewnick, G. Haug, T. Klimach, J. Sciare, H. Su, U. Pöschl

## Abstract

The role of aerosolized SARS-CoV-2 viruses in airborne transmission of COVID-19 is debated. The transmitting aerosol particles are generated through the breathing and vocalization by infectious subjects. Some authors state that this represents the dominant route of spreading, while others dismiss the option. Public health organizations generally categorize it as a secondary transmission pathway. Here we present a simple, easy-to-use spreadsheet model to estimate the infection risk for different indoor environments, constrained by published data on human aerosol emissions, SARS-CoV-2 viral loads, infective dose and other parameters. We evaluate typical indoor settings such as an office, a classroom, a choir practice room and reception/party environments. These are examples, and the reader is invited to use the algorithm for alternative situations and assumptions. Our results suggest that aerosols from highly infective subjects can effectively transmit COVID-19 in indoor environments. This “*highly infective*” category represents approximately twenty percent of the patients tested positive for SARS-CoV-2. We find that “*super infective*” subjects, representing the top five to ten percent of positive-tested ones, plus an unknown fraction of less, but still highly infective, high aerosol-emitting subjects, may cause COVID-19 clusters (>10 infections), e.g. in classrooms, during choir singing and at receptions. The highly infective ones also risk causing such events at parties, for example. In general, active room ventilation and the ubiquitous wearing of face masks (i.e. by all subjects) may reduce the individual infection risk by a factor of five to ten, similar to high-volume HEPA air filtering. The most effective mitigation measure studied is the use of high-quality masks, which can drastically reduce the indoor infection risk through aerosols.

## 1. INTRODUCTION

There is growing evidence that the spreading of coronavirus SARS-CoV-2 through aerosols, i.e. small airborne particles and microdroplets in the size range up to 5 µm diameter, is a significant transmission pathway of COVID-19.^1^ Case studies worldwide indicate that SARS-CoV-2 has viable survival rates in the air, and remains airborne for an extended period of several hours.^2^ The European Centre for Disease Prevention and Control (ECDC) states that COVID-19 is mainly transmitted via SARS-CoV-2 virus-containing respiratory droplets (i.e. larger than 5 µm in diameter), while aerosols are implicated in transmission, but the relative roles of droplets and aerosols remains unclear (https://www.ecdc.europa.eu/en/covid-19/latest-evidence/transmission). Further, the ECDC underscores the high viral load in patients close to symptom onset, suggesting that COVID-19 patients are relatively contagious at an early stage of infection. Findings for both SARS-CoV-1 and SARS-CoV-2 point to superspreading of aerosols as an important factor in the indoor disease transmission.^3,4^ This is consistent with the nearly 20 times higher likelihood of indoor compared to outdoor disease spreading.^5^ Many studies on the spread of viruses, including respiratory syncytial virus (RSV), Middle East Respiratory Syndrome coronavirus (MERS-CoV), and influenza, corroborate that viable viruses in aerosols are emitted by infected subjects, and have been detected in their environment, from which it may be concluded that people inhale the aerosolized viruses, resulting in infection and disease.^6^

Zhang and colleagues pose that airborne transmission is highly virulent and represents the dominant route in the spreading of COVID-19.^7^ They find that the wearing of face masks has been a critical aspect in the outcome of COVID-19 trends in three main areas affected by the pandemic (Wuhan, Italy, New York). Other measures, such as social distancing, appeared to have been insufficient, suggesting an important role of aerosols as they disperse over relatively large distances. Schools represent an environment of special interest, with compulsory attendance, and their closure can be societally disruptive. Nonetheless, schools have been closed, which has helped reduce the disease incidence and mortality, especially in the early stages of COVID-19 outbreaks.^8^ SARS-CoV-2 viral loads in infected children appear to be similar to those in adults.^9,10^ On the other hand, the incidence of COVID-19 under children may be less than among adults.^11,12,13^ Transmission of SARS-CoV-2 between children in schools was predicted to be less efficient than of influenza, leading to the conclusion that school closures may prevent only a small percentage (<5%) of COVID-19 deaths.^14^ Furthermore, it was articulated that by demonstrating aerosol generation by speaking and coughing, or by recovering viral RNA from the air, the aerosol-based transmission is not proven – infection also depends on the route and duration of exposure, the infection dose and host defenses.^15^ These authors also argue that infection rates and transmission in populations during normal daily life are difficult to reconcile with aerosol-based transmission.

Apparently, the role of aerosols in the spreading of SARS-CoV-2 is controversial. Here we present a simple, but transparent and easily adjustable spreadsheet algorithm to estimate the indoor infection risk from aerosolized viruses, based on adjustable parameters such as room size, number of exposed subjects, inhalation volume, and aerosol production from breathing and vocalization. We apply it to study the role of aerosol transmission of SARS-CoV-2 in indoor locations, based on assumptions about the viral load and infection dose, for example, constrained by data from the literature. We focus on aerosol transmission, hence implicitly assuming that contact infection and by larger droplets are either minimal, e.g. through social distancing and hygienic measures, or that these transmission routes should be considered additionally. Our approach aims to provide insight into the effectiveness of mitigation measures against indoor COVID-19 infection through aerosols.

## 2. METHODS

To estimate the COVID-19 infection risk from airborne transmission, we developed a spreadsheet model algorithm that includes a number of modifiable environmental factors that represent relevant physiological parameters and environmental conditions. The spreadsheet can be found in the Supporting Information, which is also available as a user-friendly, online calculation tool (https://www.mpic.de/4747065/risk-calculation), for which the parameters are listed in Table 1. The standard setting represents a case for a 60 m^2^ large and 3 m high room with 24 susceptible subjects (i.e. a total of 25 subjects), representing a classroom where one pupil is infectious for two days, being present during six hours per day. For simplicity, all subjects are assumed to be equal in terms of breathing, speaking and susceptibility to infection. The two-day period represents that of highest infectiousness of the index subject, after which s/he is assumed to either develop symptoms and stay at home or become substantially less infectious. The spreadsheet parameters can be easily adjusted to account for different environmental conditions and activities. In addition to the classroom setting, we consider three common types of environment, as indicated in Table 2. To study the impact of measures that can mitigate infection risks, we consider five scenarios (Table 3). In the next section we review the recent literature to underpin the parameter settings and ranges. We emphasize that continued progress in scientific understanding of COVID-19 may require future adjustments of assumptions and parameter settings.

**Table 1.** Spreadsheet parameters and ranges applied to compute infection risk. Standard setting refers to a classroom (indoor environment nr. 2). The spreadsheet can be downloaded from the Supplement and used as an online calculation tool: https://www.mpic.de/4747065/risk-calculation.

| Parameters | Standard | Range | Units |
| --- | --- | --- | --- |
| Infectious episode (exposure) | 2 | 0.08 – 5 | days |
| Wet aerosol diameter | 5 | 2 – 10 | µm |
| Virus lifetime in aerosol | 1.7 | 0.6 – 2.6 | hours |
| Emission during breathing | 0.1 | 0.06 – 1.0 | cm<sup>-3</sup> |
| Emission during speaking (singing) | 1.1 | 0.06 – 3.0 | cm<sup>-3</sup> |
| Speaking/breathing ratio | 0.10 | 0 – 1 | - |
| Respiratory rate | 10 | 5 – 20 | L/min |
| Viral load “highly infectious” | 5×10<sup>8</sup> | 10<sup>8</sup> – 10<sup>9</sup> | RNA copies/cm<sup>3</sup> |
| Viral load “super infectious” | 5×10<sup>9</sup> | 10<sup>9</sup> – 10<sup>10</sup> | RNA copies/cm<sup>3</sup> |
| Deposition probability in lungs | 0.5 | 0.2 – 0.8 | - |
| Infective dose (D50) | 316 | 100 – 1000 | RNA copies |
| Room area | 60 | 40 – 100 | m<sup>2</sup> |
| Room height | 3 | 3 – 4 | m |
| Subjects in room | 25 | 4 – 100 | persons |
| Passive ventilation rate | 0.35 | 0 – 1 | hour<sup>-1</sup> |
| Active ventilation rate (with outside air) | 2 | 2 – 9 | hour<sup>-1</sup> |
| Face mask filter efficiency from inhalation plus exhalation | 0.7 | 0 – 0.95 | - |

**Table 2.** Example indoor environments. There is one index subject present for the indicated time period.

| Indoor environment | Room size<br>(m<sup>2</sup>) | Room height<br>(m) | Subjects present | Exposure duration<br>(hours) |
| --- | --- | --- | --- | --- |
| 1. Office | 40 | 3 | 4 | 16 |
| 2. Classroom | 60 | 3 | 25 | 12 |
| 3. Choir practice | 100 | 4 | 25 | 3 |
| 4. Reception | 100 | 4 | 100 | 3 |

**Table 3.** Scenarios of mitigation measures. VR is ventilation rate with outside air, or by high-volume HEPA filtering (last column).

| Scenario | VR 0.35 hr<sup>-1</sup> | VR 2 hr<sup>-1</sup> | Masks, 70%<br>efficiency | Masks, 95%<br>efficiency | High-vol<br>VR 9 hr<sup>-1</sup> |
| --- | --- | --- | --- | --- | --- |
| A. Standard<br>(passive ventilation) | √ |  |  |  |  |
| B. Active ventilation |  | √ |  |  |  |
| C. Active ventilation<br>plus facial masks |  | √ | √ |  |  |
| D. Active ventilation<br>plus FFP2 masks |  | √ |  | √ |  |
| E. High volume air<br>filtration with HEPA |  |  |  |  | √ |

In the standard case for a room with closed windows, we assume a passive indoor air volume exchange rate of 0.35 hr^-1^.^16^ If active blow ventilation with outside air is applied for short periods every hour, the rate increases to 2 hr^-1^, while the application of one or more high-volume air filtering devices (with High-Efficiency Particulate – HEPA filters) presumably reduce the aerosol concentration according to an equivalent exchange rate of 9 hr^-1^. Obviously, the exchange rates can vary depending on the capacity of the devices. We emphasize that the conditions presented here exemplify a few prominent cases, and the effectiveness of the mitigation measures are mere illustrations. By modifying the parameter settings in the spreadsheet, a range of conditions, settings and mitigation measures can be simulated. For example, a respiration rate of 10 L/min (Table 1) is representative for a person between being at rest and performing light activity, whereas calculations for exercising subjects in a fitness center would have to scale this up by a factor of three to five. Further, children respire a smaller volume of air than adults but at higher frequency. Finally, we assume rapid mixing of the air in the rooms (within minutes), which seems reasonable considering the turbulent atmosphere with localized human heat sources and movement, but it imposes the restriction of room sizes up to about 100 m^2^. Although our spreadsheet algorithm is simple, it represents the available scientific knowledge of aerosol transmission, and transparently shows how different conditions, parameters and precautionary measures can influence SARS-CoV-2 infection.

## 3. PARAMETER CONSTRAINTS

### 3.1. Particle size

Humans produce infectious aerosols in a wide range of particle sizes, but viruses and other pathogens are predominately found in the small particles, which are immediately respirable by individuals who are exposed.^17^ Speech and coughing generate distinct aerosol and droplet size modes at median diameters of around 1 µm and 2 µm, and also relatively large ones of >100 µm. The modes are associated with three processes: one occurring deeply in the lower respiratory tract, another in the region of the larynx and a third in the upper respiratory tract including the oral cavity. The first contains the respiratory droplets produced during normal breathing. The second, the laryngeal mode, is most active during vocalization and coughing. The third, the oral cavity mode, is active during speech and coughing.^18^ Research on seasonal influenza transmission showed that sneezing and coughing are relatively unimportant infection pathways, and are not needed to explain virus aerosolization because of their relatively low frequency of occurrence.^19^ Therefore, we disregard coughing and sneezing as sources of infectious particles, also because symptomatic subjects should not appear in public buildings.

Typically, 80 to 90% of particles from breathing and speaking have a diameter around 1 µm, defined as aerosol particles, i.e. shortly after their emission as respiratory droplets, which dry in the environment. Upon emission from the human respiratory system at a relative humidity (RH) close to 100%, the particles are initially wet and relatively large, but in ambient air, at reduced RH, they rapidly dehydrate and transform into more-or-less dry aerosols. Aerosol particles of about 1 µm diameter evaporate in milliseconds, small droplets of 10 µm in less than a second, while relatively large droplets, with diameters >100 µm, can survive for almost a minute and fall to the ground before they evaporate.^20,21,22^ The number, size and volume of the droplets emitted according to the level of vocalization, plus the viral SARS-CoV-2 load determine the flux of viruses into the environment. The viral load in the aerosols may be equal or even higher than in the larger droplets.^17^ Based on observed number and volume size distributions of respiratory droplets emitted during breathing and speaking and the size dependence of viral load, we assume that the emitted droplets initially have an effective average diameter of 5 µm, quickly shrinking to aerosol particles to around 1 µm in ambient air.^17,18^

### 3.2. Particle emissions and vocalization

The concentrations of particles (microdroplets) produced from human speech can range from 0.1–3 cm^-3^ depending on loudness.^20^ Morawska and colleagues report a particle production of about 0.1–1.1 cm^-3^ for the range between breathing and sustained vocalization.^23^ Singing is a particularly strong aerosol source, and can emit two orders of magnitude more aerosol particles than breathing. At the quietest volume, neither singing nor speaking are significantly different from breathing.^24^ Note that the unit cm^-3^ is related to the volume respiration rate to obtain the emission flux. Some individuals are super emitters, producing aerosol particles at an order of magnitude higher rate than others, although the mechanism is unknown.^25^ Aerosol super emitters are not very common, and could be symptomatic or not. If such an infected person is engaged in loud speaking or singing, the formation of droplets and aerosols, and thus viral emissions, can increase by orders of magnitude, which may explain the occasional superspreading events in crowded situations.^20,23,26,27^ Considering the reported ranges of particle size distributions and emission rates, and an effective mean diameter of ∼5 µm as outlined above, we adopt characteristic particle number concentrations of 0.06 cm^-3^ for breathing, 0.6 cm^-3^ for speaking, and 6 cm^-3^ for singing.^17,18,20,23,25,26^

### 3.3. Viral load

The viral load of emissions from symptomatic and asymptomatic cases has not yet been directly determined, and information on the number of SARS-CoV-2 copies in the air is scarce. The working hypothesis is that the respiratory aerosols/droplets emitted by infected persons have the same viral load as found in the fluids within the airways that generate these aerosols/droplets.^17,28^ The median incubation period of COVID-19 is about 5 days.^29^ It is particularly relevant to focus on pre-symptomatic and asymptomatic transmission, as individuals with symptoms are expected to stay at home. Both have been observed, and transmission can occur 1-3 days before the development of symptoms, which may cause about half of the infections,^30^ also mentioned by the ECDC. From a literature review, it was concluded that the average fraction of SARS-CoV-2 infections that remain asymptomatic is about 20% (95% confidence interval is 17–25%).^31^ These significant fractions of infections from pre- and asymptomatic index cases provide extra motivation for mitigation measures.

The typical range of RNA copies in specimens from the throat and sputum of COVID-19 patients is 10^4^ –10^11^ mL^-1^.^9,10,11,32,33,34^ The viral load has been found to increase with advanced age and with the severity of COVID-19 outcomes.^34,35^ German and Swiss teams together collected around 80,000 specimens, of which ten to twenty percent was positive in RT-PCR tests, respectively.^10,11^ It appeared that the viral loads do not differ between age categories, but young subjects (especially under ten years) tested positive less frequently. This may be due to less frequent diagnoses among children, associated with the absence of symptoms or only mild symptoms.^10^ Infectiousness was found to be highest around the onset of symptoms.^10,11,32,34,36^

Here we focus on the phase when subjects are particularly infectious, and find that this represents a fraction of about one fifth of the positive-tested patients, during which they have a mean viral load of about 5×10^8^ mL^-1^ (i.e. within the range 10^8^–10^9^ mL^-1^). This range was found to occur somewhat more frequently in the Swiss compared to the German data. We use this estimate to define the category “*highly infectious*”. Between five and ten percent of positive-tested subjects had an even higher mean viral load of about 5×10^9^ mL^-1^, which is adopted for the category “*super infectious*” (i.e. more than 10^9^ mL^-1^). The latter category was found to occur more frequently in the German compared to the Swiss data, which may be due to the predominant testing of throat swaps specimens in the former and a large fraction of nasal swaps in the latter. We note that these values may need to be adjusted subject to ongoing studies. The corresponding input parameters can be easily replaced in the spreadsheet algorithm, but such changes are not expected to alter the conclusions about the effectiveness of the investigated precautionary measures (e.g. ventilation and face masks).

### 3.4. Virus lifetime in aerosol

Reports of so-called super spreader events corroborate that aerosols can play a role in SARS-CoV-2 transmission, because many infected persons were well away (>1.5 m) from the index subject. For example, during choir rehearsal, a single person could infect most of the group members.^37,38^ The emission was amplified by the singing. In indoor air, SARS-CoV-2 is able to remain viable for at least three hours post-aerosolization, similarly to SARS-CoV-1.^39^ The half-life of the SARS-CoV-2 virus on aerosols is about 1.1–1.2 hours within a 95% confidence interval of 0.64–2.64 hours. For MERS-CoV a similar airborne survival period was found, which declined in hot and dry air conditions.^40^ From studies of “surrogate” RNA viruses, assuming analogous behavior of SARS-CoV-2, it seems that virus survival in the air increases with decreasing temperature and relative humidity.^41^ This could be an issue in regions with cold and dry winter conditions, as ventilating with outside air will decrease both the temperature and relative humidity in the room, thus reducing the virus concentrations but increasing their lifetime. Humidity can also affect the size and lifetime of the aerosol particles.^42^ Here we neglect these hitherto unquantified influences of temperature and humidity, and adopt the half-life measured by van Doremalen and colleagues, resulting in a lifetime (e-folding time) of 1.7 hours.^39^

### 3.5. Particle deposition probability

If a safe distance of about 1.5 m is met, most of the large droplets emitted by infected subjects will settle gravitationally and tend to not reach recipients (except for coughing and sneezing).^43^ However, aerosolized viruses remain airborne, and their inhalation leads to effective uptake in the nasal fossa, pharynx and larynx, trachea and lungs.^44^ After being inhaled, particles of >20 µm diameter are predominantly deposited in the upper respiratory tract, while more than 90% of the fine aerosol particles with a diameter smaller than 2.5 µm can penetrate deeply into the lungs.^44,45^ By simulating inhalation and associated turbulent air motions in the respiratory system, it was estimated that 100 µm particles deposit in the nasal fossa, while 15 µm particles have a 62% probability to reach the lungs; and for 1 µm particles this was estimated at 94%.^44,45,46^ Since most viruses are contained in the small particles, it may be assumed that they penetrate deeply and, when retained, have the potential to cause infection in the lungs.

Measurements and model simulations of particle deposition within the respiratory system indicate that for 1 µm particles the deposited fraction is relatively low, about 0.2.^47,48^ However, this applies to hydrophobic particles studied in laboratory settings. Since the SARS-CoV-2 containing particles are hydrophylic and the relative humidity in the respiratory tract is close to 100%, it is important to account for the aerosol hygroscopic growth, which increases the inertia and sedimentation of the particles, hence a deposition probability of 0.5 was found to be more realistic.^49,50^ This was corroborated by measurements of ambient particle deposition in human subjects.^51^ It should be mentioned that cells in the conducting zone may be particularly vulnerable to SARS-CoV-2 attack (increased expression of ACE2 receptors), hence in this respect the larger aerosol particles and droplets, which penetrate less deeply, could be particularly significant.^52,53^

### 3.6. Face mask efficiency

Many studies have reported that facial masks substantially reduce the infection risk, which applies to disposable surgical masks as well as reusable cloth masks.^54,55,56,57,58,59,60^ When worn as designed, surgical and cloth masks can have filtration efficiencies of 53–75% and 28–90%, respectively.^61^ Mask wearing works in two ways, by preventing infected subjects from spreading droplets and aerosols, and by limiting exposure through inhalation. Further, homemade masks with multiple fabric layers can also effectively reduce droplet dispersal.^62^ Drewnick and colleagues investigated many materials, and measured a large range of filtration efficiencies, from about 10% to 100%.^63^ Materials with small leak areas, e.g. 1–2%, were shown to have substantially reduced efficiency. By the stacking of a number of fabric layers, homemade masks appeared to achieve good filtration efficiencies of about 50–80% for 0.5–10 µm particles, and 30–60% for 30–250 nm particles. These results agree with previous studies, and confirm that surgical masks are generally more efficient than homemade cloth products.^64,65^

The filtering efficiency of medical and non-medical masks is typically exceeded by N95 (filter 95% of the particles), FFP2 (filter ≥94%) and FFP3 (≥99%) respirators, which are recommended for health-care workers.^57,66^ The universal, public wearing of masks (not respirators) by at least 80% of the population was shown to be particularly effective in reducing the spreading of COVID-19.^67^ The effectiveness of using face masks by the general population obviously increases with their filter efficiency, but even if it is low, the benefits are apparent.^60^ By studying policies in different countries, Kai and colleagues found a strong correlation between mask wearing and both daily and peak growth reduction of COVID-19, with successful application in countries such as China, South Korea and Japan, where this practice is ubiquitous.^67^

For our purpose we assume an inhalation filtering efficiency of about 30%, and a reduction in droplet and aerosol emissions of about 60%. This yields a total risk reduction of about 70% when all subjects in the room are compliant, in accord with the significant face mask efficacy derived by Cheng and colleagues^17^. Different values can be easily tested with our algorithm, and we also consider a scenario with 95% efficiency, i.e. of a high-quality mask (Table 3). Note that the ubiquitous wearing of respirators (e.g. N95 or FFP2), which very efficiently limit the inhalation and exhalation of particles, would decrease the infection risk even more strongly by more than 99%.

### 3.7. Infective dose D50

The number of viral copies that are needed to cause an infection can vary depending on host defenses, for example. It is expressed as D50, being the mean dose that causes an infection in 50% of susceptible subjects. The D50 is not well known for COVID-19, and we have to rely on indirect information, notably because human volunteers would be needed to obtain this information. By applying a dose-response model for SARS-CoV-1, based on animal studies, a D50 of 280 viral copies was derived (and a D10 of 43 copies), which is similar to that for the common cold and animal coronaviruses.^68^ Another study mentioned that the D50 for Influenza-A is 790 viral copies, but that its infectiousness is much lower than of SARS-CoV-2.^69^ By employing the SARS-CoV-1 model to a disease outbreak in Hong Kong, a D50 between 16 and 160 viruses per person was back-calculated.^68^ A review of data and comparisons with SARS-CoV-1 and MERS concluded that the D50 for SARS-CoV-2 in humans is less than 1000 viral copies, but slightly higher than estimated for SARS-CoV-1.^70^

Here we assume that D50 is in the range of 100–1,000, and by taking the geometric mean, we arrive at D50 = 316 viral copies. This corresponds to a virus titer of about 220 plaque forming units (PFU≃0.7⋅D50, with PFU being a measure of concentration in virology).^70^ This means that the infection risk of a single viral RNA copy is:

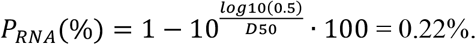

The infection risk of a individual person in the room follows from

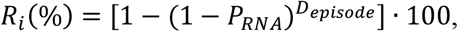

in which *D*_episode_ represents the number of viral copies inhaled and deposited in the airways. The risk of one of the persons in the room being infected is

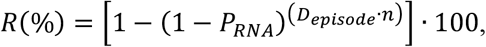

where *n* is the number of susceptible subjects in the room. Note that the number of inhaled viral copies follows a Poisson distribution, but because viruses are considered to have independent and identical infectivity, the calculated infection probability is the same regardless of whether a Poisson distribution is considered or not.

## 4. RESULTS

In our assessment of indoor infection risks and the effectiveness of mitigation measures, we consider a single infected person (index case) present within the group in each of the four investigated example environments. This person is likely to be pre-symptomatic (typically 1–3 days), as the highest viral load mostly occurs around the onset of symptoms, after which the patient is expected to stay at home. Alternatively, the index subject can be asymptomatic, which happens in about 20% of cases, although it is not clear whether such individuals are equally infectious. Figure 1, and Tables 4 and 5 present the risk assessment for standard conditions (highly infectious index case) and super spreader events, respectively. For the latter we assume that either the viral load is a factor of ten higher than for the highly infectious index subject (thus being super infectious) or the person is a super aerosol-emitter. It is theoretically possible that both occur in the same person, but the incidence of such cases is probably very low, hence we disregard this option.

**Table 4.** COVID-19 infection risk in four example environments for five scenarios. Unit: percent per episode. The individual risk is that of a particular person being infected, the group risk refers to the infection risk of at least one person in the group.

| **Environment** | **Scenario A**<br>VR 0.35 hr<sup>-1</sup> | **Scenario B**<br>VR 2 hr<sup>-1</sup> | **Scenario C**<br>Masks, 70%<br>efficiency | **Scenario D**<br>Masks, 95%<br>efficiency | **Scenario E**<br>High-vol<br>VR 9 hr<sup>-1</sup> |
| --- | --- | --- | --- | --- | --- |
| **1.1. Office**<br>Individual risk | 18 | 7.0 | 2.2 | 0.4 | 1.9 |
| **1.2. Office.**<br>Group risk | 45 | 20 | 6.3 | 1.1 | 5.7 |
| **2.1. Classroom**<br>Individual risk (>10 yr) | 9.5 | 3.6 | 1.1 | 0.2 | 1.0 |
| **2.2. Classroom**<br>Group risk (>10 yr) | 91 | 58 | 23 | 4.3 | 21 |
| **2.3. Classroom**<br>Individual risk (≤10 yr) | 1.0 | 0.4 | 0.1 | 0 | 0.1 |
| **2.4. Classroom**<br>Group risk (≤10 yr) | 21 | 8.3 | 2.6 | 0.4 | 2.3 |
| **3.1. Choir practice**<br>Individual risk | 29 | 12 | - | - | 3.3 |
| **3.2. Choir practice**<br>Group risk | 100 | 95 | - | - | 55 |
| **4.1. Reception**<br>Individual risk | 4.1 | 1.5 | 0.5 | 0.1 | 0.4 |
| **4.2. Reception**<br>Group risk | 98 | 78 | 36 | 7.2 | 33 |
| **4.3 Party**<br>Individual risk | 27 | 11 | 3.4 | 0.6 | 3.0 |
| **4.4. Party**<br>Group risk | 100 | 100 | 97 | 43 | 95 |

**Table 5.** COVID-19 infection risk in four example environments for five scenarios. The index subject is assumed to be either super infectious (10× higher viral load) or a super emitter (10× higher aerosol emission rate). Unit: percent per episode. The individual risk is that of a particular person being infected, the group risk refers to the infection risk of at least one person in the group.

| **Environment** | **Scenario A**<br>VR 0.35 hr<sup>-1</sup> | **Scenario B**<br>VR 2 hr<sup>-1</sup> | **Scenario C**<br>Masks, 70%<br>efficiency | **Scenario D**<br>Masks, 95%<br>efficiency | **Scenario E**<br>High-vol<br>VR 9 hr<sup>-1</sup> |
| --- | --- | --- | --- | --- | --- |
| **1.1. Office**<br>Individual risk | 87 | 52 | 20 | 3.6 | 18 |
| **1.2. Office**<br>Group risk | 100 | 89 | 48 | 10 | 44 |
| **2.1. Classroom**<br>Individual risk (>10 yr) | 63 | 30 | 10 | 1.8 | 9.3 |
| **2.2. Classroom**<br>Group risk (>10 yr) | 100 | 100 | 93 | 35.3 | 90 |
| **3.1. Choir practice**<br>Individual risk | 97 | 71 | - | - | 29 |
| **3.2. Choir practice**<br>Group risk | 100 | 100 | - | - | 100 |
| **4.1. Reception**<br>Individual risk | 34 | 14 | 4.5 | 0.8 | 4.0 |
| **4.2. Reception**<br>Group risk | 100 | 100 | 99 | 53 | 98 |

**Figure 1.**
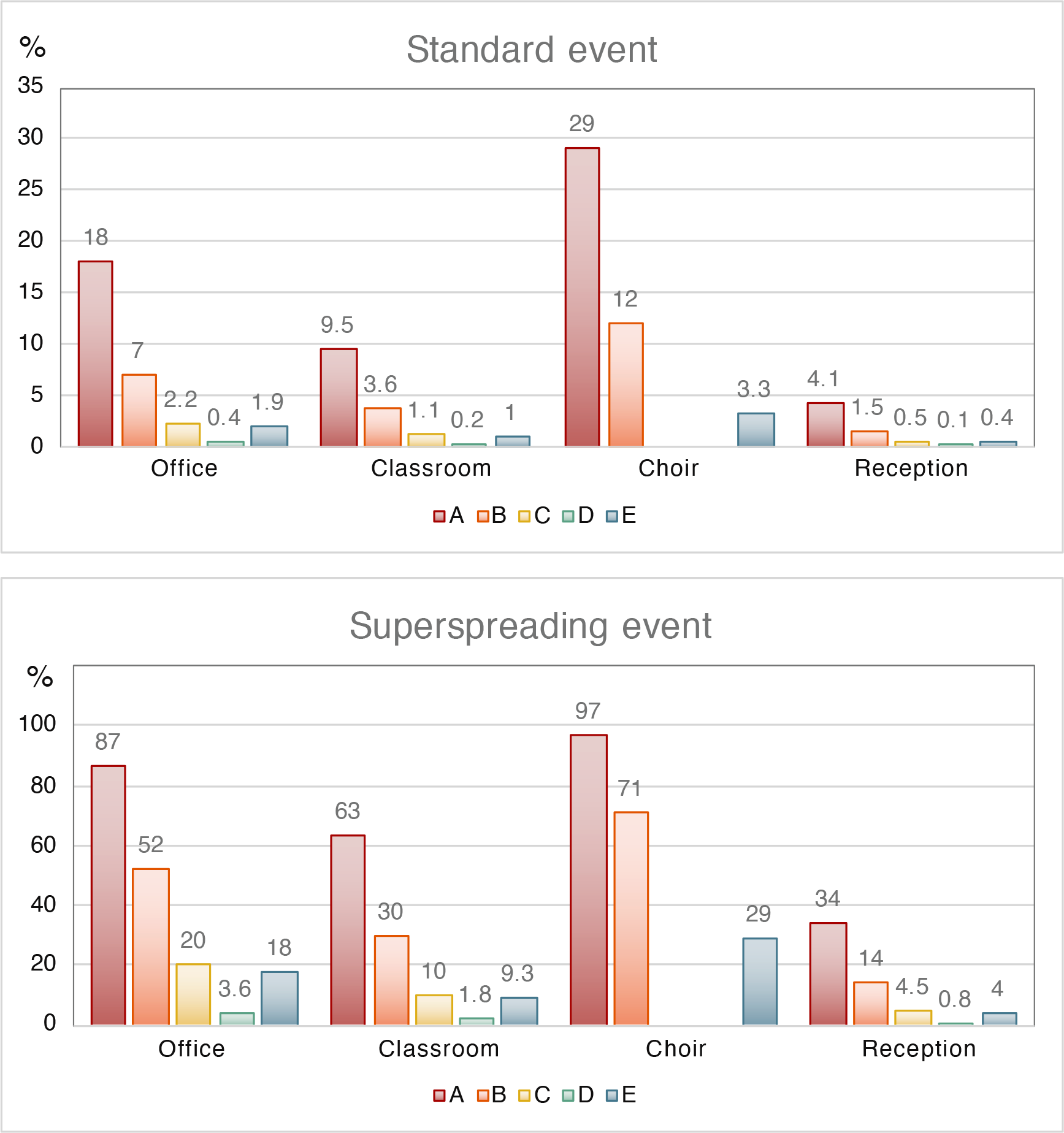
Individual risk of a particular person being infected (equivalent to the fraction of the group being infected) in four indoor environments and five scenarios, for standard and superspreading conditions. Scenario A: passive ventilation, no masks. Scenario B: active ventilation with outside air, no masks. Scenario C: active ventilation, facial masks (not for choir). Scenario D: active ventilation, high-quality masks (not for choir). Scenario E: High-volume filtration with HEPA.

### 4.1. Office environment

Here we assume the presence of four subjects, one being highly infectious for two days (eight hours/day). Under scenario A (only passive ventilation), the risk that one of the others becomes infected is 45%, and the individual infection risk is 18%. Active ventilation with outside air (B) reduces these risks by a factor of 2 to 2.5, and by additionally wearing face masks (C) the risks are reduced by a factor of 7 to 8 (Table 4). A similar risk reduction (factor of 8 to 9) is achieved by applying high-volume HEPA filtering (E). The wearing of high-quality masks (D) leads to the largest risk reduction by more than a factor of 40 (i.e. that one of the others becomes infected). In the case of a super spreader event, two to three colleagues would likely be infected, which could be reduced to one to two by active ventilation (B). An overall reduction of the individual risk by a factor of 4 to 5 could be achieved by additionally wearing masks or by high-volume HEPA ventilation (C, E). The individual risk could be reduced by a factor of 25 by wearing high-quality masks (D). Although these conditions refer to an office environment, the risk reduction achieved by active ventilation and wearing masks also applies to other environments with similar dimensions and numbers of subjects, for example a hospital ward or a room in a nursing home.

### 4.2. Classroom environment

We putatively distinguish the young (children under ten years) from the older pupils, because especially the young ones may potentially be less infective (or less often infective), for which we assume a reduced viral load by a factor of ten. Our results in Table 4 show that the risk of COVID-19 from a reduced-infective subject in this youngest group is about 21% (scenario A), while the individual risk is about 1%. For the older pupils with a highly infective index subject this is 91% and nearly 10%, respectively. Ventilating with outside air is helpful, as it reduces the individual aerosol infection risk for the ones under ten to a very low fraction, i.e. well under a percent, assuming that these children are present during two days (six hours/day), and perhaps even less as the daily time in the classroom could be shorter than six hours. For the older children, active ventilation with outside air will reduce the risk of one becoming infected to about 58% (B). A low individual risk of about 1% may be achieved for these children by additionally wearing face masks (C).

In a classroom-type environment with sensitive subjects (i.e. with enhanced risk of severe COVID-19 progression), high-volume HEPA filtration could be considered to reduce the individual infection risk by a factor of 9 to 10, or more when combined with other mitigation measures. This would work for classrooms in general, of course, but expensive filtration techniques should probably be reserved for those who need it most. In the case of a super spreader among the pupils or teacher, the risk is high, and 63% of the others could be infected. Even with active ventilation and mask wearing it seems unavoidable that a few will be infected, but these measures can nevertheless decrease the individual infection risk substantially, i.e. by a factor of 6 to 7 (Table 5). High-quality masks (or even better respirators) could keep the infection risk in check, but it may not be realistic to expect this in schools (as it is for nursing staff). Note that we do not distinguish between pupils and the teacher, who is likely to do much of the talking, associated with significant particle emissions, which could be accounted for by refining the settings in the spreadsheet.

### 4.3. Choir practice

The infection risk for choir members is clearly enhanced, in spite of the twofold larger room volume compared to the classroom with the same number of subjects present (Table 4, Figure 1). This is due to the much higher rate of aerosol emission during singing compared to breathing, for which we assume a breathing/vocalization ratio of 0.25 and a concentration of 6 cm^-3^ for the singing, and a mean respiratory rate of 15 L/min during the event with a duration of three hours. This respiratory rate is higher than of trained but lower than of untrained singers.^71^ We can compare with an actual COVID-19 infection event during a choir rehearsal in the state of Washington in the USA on 10 March 2020, for which conditions are well-documented.^38^ Our algorithm reproduces the observed very high infection rate (>80%) for a super infectious subject in the room and a breathing/vocalization ratio of about 0.3, very close to the algorithm settings applied here. For the results presented in Figure 1 and Table 4, we again assumed one highly infective index case in the choir, attaining an infection risk of 29% (scenario A). This can be reduced by more than a factor of two through active ventilation (B) and nearly a factor of nine by high-volume HEPA filtering of the air (E). We do not consider face masks and respirators for singing practice (C and D). In the event of a super spreader in the room, the infection risk is very high, and even with active ventilation or high-volume HEPA filtering, it is 71% (B) or 29% (E), respectively (Table 5, Figure 1).

### 4.4. Reception

For the reception guests we assumed slightly louder speaking than in the office and classroom, leading to a particle concentration of 1.0 cm^-3^, a speaking/breathing ratio of 0.25 and a respiration rate of 12 L/min. We obtain a lower individual infection risk than for the school class, because the room is bigger and the duration of exposure is shorter. However, in the reception room many more susceptible subjects are present, which scales to the number that may be infected by one index case, being about four in the absence of mitigation measures, compared to about two in the classroom (Scenario A). The reception would become less risky with active ventilation and by wearing masks, and especially by highly efficient masks (C, D). By applying active ventilation with outside air, the individual risk is reduced by a factor of 2.5 to 3, while high-volume HEPA filtering decreases it by a factor of ten (Table 4). When the index case is a super spreader, the risk of a large number of infections is high under scenario A without mitigation (34%), which could be reduced to 14% (B) or below 9% (E), with active ventilation or high-volume HEPA filtering, respectively (Table 5). The risk that four or more guests will become infected is significant under all scenarios, except when all would wear high-efficiency masks.

### 4.5. Cluster infections

Figure 1 illustrates that the individual infection risk from one index subject in the office, classroom and during the reception decrease in this order by factors of about two, and that during choir practice risks are relatively high. Note that the individual infection risk equals the fraction of the group that is at risk, thus being increasingly significant with the number of subjects present. If one index case impacts a large group of subjects there is a risk that COVID-19 clusters (>10 infections) develop, which need to be contained swiftly, which can be expected from superspreading events. In the present examples, the superspreading risk for the classroom would be about 15 infection cases, 23 during choir practice and 34 at the reception. Hitherto we have assumed that the reception guests are not singing nor speaking loudly (i.e. a quiet and orderly reception without too many speeches), which is unlikely to hold true when alcohol is consumed and musical entertainment is offered with dancing. This qualifies as a party, which appears to be relatively risky, and a cluster infection could even occur without a super spreader (Table 4, Figure 2). Many guests could become infected under scenario A (27%). Since party guests may not be interested in wearing face masks, the high-volume HEPA solution could be considered, which would reduce this number by a factor of nine, i.e. to well below the size of a cluster (about 3 cases). Such risk reduction is typical for high-volume HEPA filtering (capacity should be adapted to the room size).

**Figure 2.**
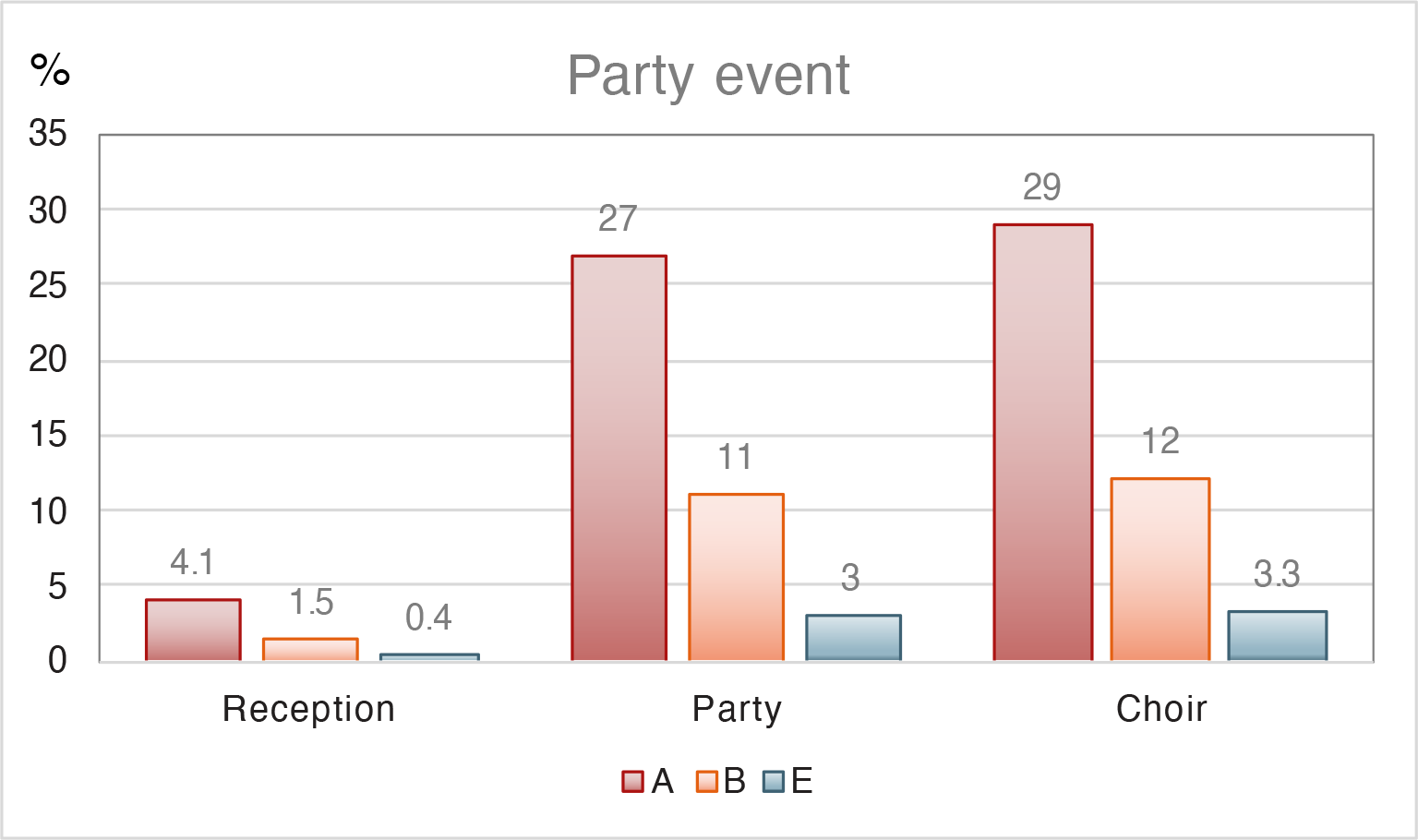
Individual risk of a particular person being infected (equivalent to the fraction of the group being infected), comparing a party with a reception and a choir practice, for three scenarios. Scenario A: passive ventilation, no masks. Scenario B: active ventilation with outside air, no masks. Scenario E: High-volume filtration with HEPA.

## 5. DISCUSSION

There are significant uncertainties that may influence the results of our calculations, as outlined in Section 3 where we presented the parameter constraints. Important uncertainties are related to the infective dose and the viral load, which may particularly apply to the risk estimates of children. Asymptomatic SARS-CoV-2 subjects may carry the same viral load as symptomatic ones, but the available data do not allow firm conclusions. Buonanno and colleagues affirm that high emission rates can be reached by asymptomatic, infectious SARS-CoV-2 subjects performing vocalization during light activities, whereas a symptomatic subject in resting conditions typically has a one to two order of magnitude lower emission rate.^72^ In schools, teachers may speak forcefully, and especially young children can be loud, which will enhance the spreading. In households, infections are higher from symptomatic than asymptomatic subjects. Since symptomatic individuals are expected to stay at home and avoid public places, the pre-symptomatic, asymptomatic and mild COVID-19 cases (the latter possibly confused with a common cold) pose the largest infection risk in indoor environments.^27,59^

To account for uncertainties and variability among the population, we considered a range of conditions, e.g. from small children with putatively lower viral loads up to super spreaders. Even though the estimated infection risks vary with basic assumptions, associated with uncertainty, the predicted reductions from different mitigation measures such as active ventilation with outside air, the ubiquitous wearing of face masks of intermediate and high quality, and air filtering are robust. Of course, they can be optimized, e.g. through the properties of masks and the rates of air exchange. Our sensitivity calculations for 95% efficient, high-quality masks show that much can be gained by paying attention to the mask properties and how they are worn. It is evident that SARS-CoV-2 can be transmitted through the air by aerosol particles, causing COVID-19 infections in the indoor environment. But uncertainties remain significant, while additional scientific research will need time to quantify infection risks and pathways in greater detail. However, by waiting for additional scientific results, valuable time will be lost that could be used to control the pandemic.^73^

Obviously, poor ventilation increases the time period during which aerosols accumulate in the room. Risks can be significantly reduced by regular ventilation with outside air and by the ubiquitous wearing of face masks. As a co-benefit, this will also decrease the risk of infection with influenza and common colds, and increase student performance, for example.^74^ Multiple studies have concluded that masks help prevent or reduce virus transmission (Section 3.6). Surgical masks are relatively efficient in this respect, but also home-made cloth masks can contribute, albeit with lower efficiency. Drewnick and colleagues reported that avoiding leaks in masks is particularly important to have good overall efficacy.^63^ When masks with a sufficient number of layers of good material (especially fluffy textiles) and with proper fit are used, there could be substantial inhalation protection. The masks reduce SARS-CoV-2 emissions in both droplets and aerosol particles. At emission, the particles are initially wet (small droplets), likely remaining so under the mask. The filtration efficiency of larger droplets increases with their velocity, e.g. during coughing and sneezing.^63^ Since the face velocity and the particle size from speaking may be larger than from breathing, enhanced filtration efficiency is expected.

From SARS-CoV-2 measurements in hospitals it follows that room ventilation, open space and sanitization can effectively limit aerosol concentrations.^75^ Nevertheless, when there is a highly infectious subject in the room, especially for a prolonged period, strict respiratory protection measures are needed. In such environments, loose-fitting masks will not prevent infection, and one should consider respirators that are designed to protect the wearer, e.g. those with FFP2 and FFP3 quality. Further, high-volume air filtration systems (with HEPA filters) can help remove aerosol particles from the indoor environment. The overall efficiency is determined by the volume that can be purified per hour. This is mostly a flow-technical question and should involve characterization of air turbulence, mixing and ventilation. It was estimated that an air filtering system with a capacity up to 1,500 m^3^/hour can effectively reduce the aerosol concentration throughout a room of 80 m^2^ in a short period of time, although this calculation did not account for continuous emissions of particles.^76^

## 6. CONCLUSIONS

Based on data from RT-PCR tests in Germany and Switzerland,^10,11^ encompassing around 8,000 subjects who were confirmed as SARS-CoV-2 infected, it was found that in the most infectious phase, probably around the onset of symptoms, patients carry a mean viral load around 5×10^8^ viral (RNA) copies mL^-1^. This occurred in about one out of five of the positive tests, and we define this category as “*highly infectious*”. About one out of five-to-ten of the positive RT-PCR tests indicated an order of magnitude higher viral load, i.e. around 5×10^9^ mL^-1^, which we categorize as “*super infectious*”. Further, the super infectious category additionally includes a group of highly infectious subjects who are super aerosol-emitters, who produce an order of magnitude more particles during breathing and vocalization, although we do not have information on their frequency of occurrence. The presence of such index subjects in the room (super spreaders) is associated with a very high infection risk from aerosol transmission. We find that both categories can significantly contribute to COVID-19 risk though aerosols. It is possible that school children, especially under ten years of age, are less frequently infective. However, since school class environments are much more common than the others considered here, the overall incidence of infection outbreaks may nevertheless be high.

Super spreading events from a single index subject can cause COVID-19 clusters (>10 infections), also in classrooms. The risk of clusters from super spreaders is particularly high during choir practice (e.g. most singers infected) and on receptions (e.g. a third of the guests infected). Even with an index subject who is highly infectious (i.e. ten times less infectious than a super spreader), during party events the risk of a COVID-19 cluster is significant (e.g. 25–30 of 100 guests infected). Generally, the risk of clusters may be curbed by high-volume HEPA filtering. But since these techniques are in limited supply and costly, active ventilation and the wearing of face masks may be preferred options, as they achieve similar results at greatly reduced cost. The best, low-cost solution is the ubiquitous wearing of high-quality masks, which is estimated to be about five times more effective. But it seems unplausible that such measures will be adopted by choir singers and party guests.

We reiterate that we merely present examples, and deviations for individual environments and conditions are imminent. To assess the risk reduction from the wearing of face masks, we adopted two levels of filtering efficiency, but did not consider the large range of available types. Clearly, improved quality masks, optimal fitting and hygienic discipline will greatly increase the efficiency. Notwithstanding the uncertainties, we are confident that the relative reductions predicted for different mitigation measures such as active ventilation with outside air, the ubiquitous wearing of face masks of intermediate and high quality, and air filtering are robust. The Supporting Information offers our algorithm, which can be adapted to other environments and scenarios, under the premise that the room size is not too large (>100 m^2^), for which our rapid air mixing assumption would be violated. A www-based tool is also available (https://www.mpic.de/4747065/risk-calculation. We endorse the reasoning of Kai and colleagues,^67^ who conclude that a “mouth-and-nose lockdown is far more sustainable than a full lockdown, from economic, social, and mental health standpoints”.

## Supporting information

Spreadsheet to calculate infection risk

## Data Availability

All data used in the analysis are provided in the manuscript and supplementary materials.

## REFERENCES

(1) Cai, J., Sun, W., Huang, J., Gamber, M., Wu, J., He, G. Indirect Virus Transmission in Cluster of COVID-19 Cases, Wenzhou, China. Emerg. Infect. Dis. 2020, 26, 1343.

(2) Jayaweera, M., Perera, H., Gunawardana, B., Manatunge, J. Transmission of COVID-19 virus by droplets and aerosols: A critical review on the unresolved dichotomy. Environ. Res. 2020, 188, 109819.

(3) Lloyd-Smith, J. O., Schreiber, S. J., Kopp, P. E., Getz, W. M. Superspreading and the effect of individual variation on disease emergence. Nature 2005, 438, 355–359.

(4) Tellier, R., Li, Y., Cowling, B. J., Tang, J. W. Recognition of aerosol transmission of infectious agents: a commentary. BMC Infect. Dis. 2019, 19, 101.

(5) Nishiura, H., Oshitani, H., Kobayashi, T., Saito, T., Sunagawa, T., Matsui, T., Wakita, T., MHLW COVID-19 Response Team; Suzuki, M. Closed environments facilitate secondary transmission of coronavirus disease 2019 (COVID-19). medRxiv. 2020, https://doi.org/10.1101/2020.02.28.20029272.

(6) Morawska, L., Milton, D. K. It is time to address airborne transmission of COVID-19. Clin. Infect. Dis. 2020, ciaa939, https://doi:10.1093/cid/ciaa939.

(7) Zhang, R., Li, Y., Zhang, A. L., Wang, Y., Molina, M. J. Identifying airborne transmission as the dominant route for the spread of COVID-19. PNAS 2020, 117, 14857–14863.

(8) Auger, K. A., Shah, S. S., Richardson, T., Hartley, D., Hall, M., Warniment, A., Timmons, K., Bosse, D., Ferris, S. A., Brady, P. W., Schondelmeyer, A. C., Thomson, J. E. Association between statewide school closure and COVID-19 incidence and mortality. JAMA 2020, 324, 859–870.

(9) Wölfel, R., Corman, V. M., Guggemos, W., Seilmaier, M., Zange, S., Müller, M. A., Niemeyer, D., Jones, T. C., Vollma, P., Rothe, C., et al. Virological assessment of hospitalized patients with COVID-2019. Nature 2020, 581, 465–469.

(10) Jones, T. C., Mühlemann, B., Veith, T., Zuchowski, M., Hofmann, J., Stein, A., Edelmann, A., Corman, V. M., Drosten, C. An analysis of SARS-CoV-2 viral load by patient age. medRxiv. 2020, https://doi.org/10.1101/2020.06.08.20125484.

(11) Jacot, D., Greub, G., Jaton, K., Opota, O. Viral load of SARS-1 CoV-2 across patients and compared to other respiratory viruses. Microbes and Infection 2020, https://doi.org/10.1016/j.micinf.2020.08.004.

(12) Davies, N. G., Klepac, P., Liu, Y., Prem, K., Jit, M., CMMID COVID-19 working group; Eggo, R.M. Age-dependent effects in the transmission and control of COVID-19 epidemics. Nat Medicine 2020, 26, 1205–1211.

(13) Mizumoto, K., Omori, R., Nishiura, H. Age specificity of cases and attack rate of novel coronavirus disease (COVID-19.). medRxiv. 2020, https://doi.org/10.1101/2020.03.09.20033142.

(14) Viner, R. M., Russell, S. J., Croker, H., Packer, J., Ward, J., Stansfield, C., Mytton, O., Bonell, C., Booy, R. School closure and management practices during coronavirus outbreaks including COVID-19: a rapid systematic review. Lancet Child Adolesc. Health 2020, 4, 397–404.

(15) Klompas, M., Baker, M. A., Rhee, C., Airborne Transmission of SARS-CoV-2 Theoretical Considerations and Available Evidence. JAMA 2020, 324, 441–442.

(16) Howard-Reed, C., Wallace, L. A., Ott, W. R. The effect of opening windows on air change rates in two homes. J. Air Waste Manag. Assoc. 2011, 52, 147–159.

(17) Cheng, Y., Ma, N., Witt, C., Rapp, S., Wild, P., Andreae, M. O., Pöschl, U., Su, H. Distinct regimes of particle and virus abundance explain face mask efficacy for COVID-19. medRxiv. 2020, https://doi.org/10.1101/2020.09.10.20190348.

(18) Johnson, G. R., Morawska, L., Ristovski, Z. D., Hargreaves, M., Mengersen, K., Chao, C. Y. H., Wan, M. P., Li, Y., Xie, X., Katoshevski, D., Corbett, S. Modality of human expired aerosol size distributions. J. Aerosol Sci. 2011, 42, 839–851.

(19) Yan, J., Grantham, M., Pantelic, J., de Mesquita, P. J. B., Albert, B., Liu, F., Ehrman, S., Milton, D. K., EMIT Consortium. Infectious virus in exhaled breath of symptomatic seasonal influenza cases from a college community. PNAS 2018, 115, 1081–1086.

(20) Asadi, S., Wexler, A. S., Cappa, C. D., Santiago Barreda, S., Bouvier, N. M., Ristenpart, W. D. Aerosol emission and superemission during human speech increase with voice loudness. Sci. Rep. 2019, 9, 2348.

(21) Borak, J. Airborne transmission of COVID-19. Occup. Med. 2020, 70, 297–299.

(22) Vuorinen, V., Aarnio, M., Alava, M., Alopaeus, V., Atanasova, N., Auvinen, M., Balasubramanian, N., Bordbar, H., Erasto, P., Grande, R., et al. Modelling aerosol transport and virus exposure with numerical simulations in relation to SARS-CoV-2 transmission by inhalation indoors. Saf. Sci. 2020, 130, 104866.

(23) Morawska. L., Johnson, G. R., Ristovski, Z. D., Hargreaves, M., Mengersen, K., Corbett, S., Chao, C. Y. H., Li, Y., Katoshevski, D. Size distribution and sites of origin of droplets expelled from the human respiratory tract during expiratory activities. J. Aerosol Sci. 2009, 40, 256–269.

(24) Gregson, F. K. A., Watson, N. A., Orton, C. M., Haddrell, A. E., McCarthy, L. P., Finnie, T. J. R., Gent, N., Donaldson, G. C., Shah, P. L., Calder, J. D., et al. Comparing the Respirable Aerosol Concentrations and 1 Particle Size Distributions Generated by Singing, 2 Speaking and Breathing. ChemRxiv. 2020, https://doi.org/10.26434/chemrxiv.12789221.v1.

(25) Asadi, S., Wexler, A. S; Cappa, C. D., Barreda, S., Bouvier, N. M., Ristenpart, W. D., Effect of voicing and articulation manner on aerosol particle emission during human speech. PLoS ONE 2020, 15, e0227699.

(26) Mürbe, D., Fleischer, M., Lange, J., Rotheudt, H., Kriegel, M. Aerosol emission is increased in professional singing. 2020, 10.14279/depositonce-10374.

(27) Riediker, M., Tsai, D.-H. Estimation of Viral Aerosol Emissions From Simulated Individuals With Asymptomatic to Moderate Coronavirus Disease 2019. JAMA Netw Open. 2020, 3(7):e2013807.

(28) Fernandez, M. O., Thomas, R. J., Garton, N. J., Hudson, A., Haddrell, A., Reid, J. R. Assessing the airborne survival of bacteria in populations of 10 aerosol droplets with a novel technology. J. Roy. Soc. Int. 2019, 16, 20180779.

(29) Lauer, S. A., Grantz, K. H., Bin, Q., Jones, F. K., Zheng, Q., Meredith, H. R., Azman, A. S., Reich, N. G., Lessler, J. The Incubation Period of Coronavirus Disease 2019 (COVID-19) From publicly reported confirmed cases: Estimation and application. Ann. Intern. Med. 2020, 172, 577–582.

(30) Arons, M. M., Hatfield, K. M., Reddy, S. C., Kimball, A., James, A., Jacobs, J. R., Taylor, J., Spicer, K., Bardossy, A. C., Oakley, L. P., et al. For the Public Health–Seattle and King County and CDC COVID-19 Investigation Team 2020. Presymptomatic SARS-CoV-2 Infections and Transmission in a Skilled Nursing Facility. N. Engl. J. Med. 2020, 382, 2081–90.

(31) Buitrago-Garcia, D., Egli-Gany, D., Counotte, M. J., Hossmann, S., Imeri, H., Ipekci, M. A., Salanti, G., Low, N. Asymptomatic SARS-CoV-2 infections: a living systematic review and 1 meta-analysis. medRxiv. 2020, https://doi.org/10.1101/2020.04.25.20079103.

(32) Pan, Y., Zhang, D., Yang, P., Poon, L. L. M., Wang, Q. Viral load of SARS-CoV-2 in clinical samples. Lancet Infect. Dis. 2020, 20, 411–412.

(33) Pujadas, E., Chaudhry, F., McBride, R., Richter, F., Zhao, S., Wajnberg, A., Nadkarni, G., Glicksberg, B. S., Houldsworth, J., Cordon-Card, C. SARS-CoV-2 viral load predicts COVID-19 mortality. Lancet Respir. Med. 2020, 8, https://doi.org/10.1101/2020.06.11.20128934.

(34) To, K. K.-W., Tsang, O. T.-Y., Leung, W.-S., Tam, A. R., Wu, T.-C., Lung, D. C., Yip, C. C.-Y., Cai, J.-P., Chan, J. M.-C., Chik, T. S.-H., et al. Temporal profiles of viral load in posterior oropharyngeal saliva samples and serum antibody responses during infection by SARS-CoV-2: an observational cohort study. Lancet Infect. Dis. 2020, 20, 565–574.

(35) Yu, X., Sun, S., Shi, Y., Wang, H., Zhao, R., Sheng, J. SARS-CoV-2 viral load in sputum correlates with risk of COVID-19 progression. Crit. Care 2020, 24, 170.

(36) He, X., Lau, E. H. Y., Wu, P., Deng, X., Wang, J., Hao, X., Lau, Y. C., Wong, J. Y., Guan, Y., Tan, X., et al. Temporal dynamics in viral shedding and transmissibility of COVID-19. Nat. Med. 2020, 26, 672–675.

(37) Hamner, L., Dubbel, P., Capron, I., Ross, A., Jordan, A., Lee, J., Lynn, J., Ball, A., Narwal, S., Russell, S., et al. High SARS-CoV-2 Attack Rate Following Exposure at a Choir Practice — Skagit County, Washington, March 2020. MMWR 2020, 69, 606–610.

(38) Miller; S. L., Nazaroff, W. W., Jimenez, J. L., Boerstra, A., Buonanno, G., Dancer, S. J., Kurnitski, J., Marr, L.C., Morawska L., Noakes, C. Transmission of SARS-CoV-2 by inhalation of respiratory aerosol in the Skagit Valley Chorale superspreading event. medRxiv. 2020, https://doi.org/10.1101/2020.06.15.20132027.

(39) van Doremalen, N., Bushmaker, T., Morris, D. H., Holbrook, M. G., Gamble, A., Williamson, B. N., Tamin, A., Harcourt, J. L., Thornburg, N. J., Gerber, S. I., et al. Aerosol and surface stability of SARS-CoV-2 as compared with SARS-CoV-1. N. Engl. J. Med. 2020, 382, 1564–1567.

(40) Pyankov, O. V., Bodnev, S. A., Pyankova, O., Agranovski, I. E. Survival of aerosolized coronavirus in the ambient air. Aerosol Sci. Technol. 2018, 115, 158–163.

(41) Casanova, L. M., Jeon, S., Rutala, W. A., Weber, D. J., Sobsey, M. D. Effects of air temperature and relative humidity on coronavirus survival on surfaces. Appl. Environ. Microbiol. 2010, 76, 2712–2717.

(42) Wang, B., Wu, H., Wan, X.-F. Transport and fate of human expiratory droplets—A modeling approach. Phys. Fluids. 2020, 32, 083307.

(43) Liu, L., Wei, J., Li, Y., Ooi, A. Evaporation and dispersion of respiratory droplets from coughing. Indoor Air 2017, 27, 179–190.

(44) de Gabory, L., Alharbia, A., Kérimiana, M., Lafonc, M.-E. The influenza virus, SARS-CoV-2, and the airways: Clarification for the otorhinolaryngologist. Eur. Ann. Otorhinolaryngol. Head Neck Dis. 2020, 137, 291–296.

(45) Inthavong, K., Ge, Q. J., Li, X. D., Tu, J. T. Detailed predictions of particle aspiration affected by respiratory inhalation and airflow. Atmos. Environ. 2012, 62, 107–117.

(46) Dong, J., Shang, Y., Tian, L., Inthavong, K., Qiu, D., Tu, J. Ultrafine particle deposition in a realistic human airway at multiple inhalation scenarios. Int. J Numer. Meth. Biomed. Engng. 2019, 35, 3215.

(47) Stahlhofen, W., Rudolf, G., James, A. C. Intercomparison of experimental regional aerosol deposition data. J. Aerosol Med. 1989, 2, 285–308.

(48) Asgharian, B., Hofmann, W., Bergmann, R. Particle deposition in a multiple-path model of the human lung. Aer. Sci. Technol. 2010, 34, 332–339.

(49) Heyder, J. Deposition of inhaled particles in the human respiratory tract and consequences for regional targeting in respiratory drug delivery. Proc. Am. Thorac. Soc. 2004, 1, 315–320.

(50) Tsuda, A., Henry, F. S., Butler, J. P. Particle transport and deposition: basic physics of particle kinetics. Compr. Physiol. 2015, 3, 1437–1471.

(51) Montoya, L. D., Lawrence, J., Krishna Murthy, G. G., Sarnat, J. A., Godleski, J. J., Koutrakis, P. Continuous measurements of ambient particle deposition in human subjects. Aerosol Sci. Technol. 2004, 38, 980–990.

(52) Hsiao, T.-C., Chuang, H.-C., Griffith, S. M., Chen, S.-J., Young, L. H. COVID-19: An aerosol’s point of view from expiration to transmission to viral-mechanism. Aerosol Air Quality Res. 2020, 20, 905–910.

(53) Lukassen, S., Chua, R. L., Trefzer, T., Kahn, N. C., Schneider, M. A., Muley, T., Winter, H., Meister, M., Veith, C., Boots, A. W., et al. SARS-CoV-2 receptor ACE2 and TMPRSS2 are primarily expressed in bronchial transient secretory cells EMBO J. 2020, 39:e105114.

(54) Chu, D. K., Akl, E. A., Duda, S., Solo, K., Yaacoub, S., Schünemann, H. J. Physical distancing, face masks, and eye protection to prevent person-to-person transmission of SARS-CoV-2 and COVID-19: a systematic review and meta-analysis. Lancet 2020, 395, 1973–87.

(55) Esposito, S., Principi, N., Leung, C. C., Migliori, G. B. Universal use of face masks for success against COVID-19: evidence and implications for prevention policies. Eur. Respir. J. 2020, 55, 2001260.

(56) Fischer, E. P., Fischer, M. C., Grass, D., Henrion, I., Warren, W. S., Westman, E. Low-cost measurement of facemask efficacy for filtering expelled droplets during speech. Sci. Adv. 2020, 6, eabd3083.

(57) Howard, J., Huang, A., Li, Z., Tufekci, Z., Zdimal, V., van der Westhuizen, H., von Delft, A., Price, A., Fridman, L., Tang, L., et al. Face Masks Against COVID-19: An Evidence Review. Preprints 2020, 2020040203, https://doi:10.20944/preprints202004.0203.v1.

(58) Leung, N. H. L., Chu, D. K. W., Shiu, E. Y. C., Chan K.-H., McDevitt, J. J., Hau, B. J. P., Yen, H.-L., Li, Y., Ip, D. K. M., Peiris, J. S. M., et al. Respiratory virus shedding in exhaled breath and efficacy of face masks. Nat. Med. 2020, 26, 676–680.

(59) MacIntyre, C. R., Chughtai, A. A. A rapid systematic review of the efficacy of face masks and respirators against coronaviruses and other respiratory transmissible viruses for the community, healthcare workers and sick patients. Int. J. Nurs Stud. 2020, 108, 103629.

(60) Worby, C. J., Chang, H.-H. Face mask use in the general population and optimal resource allocation during the COVID-19 pandemic. Nat. Commun. 2020, 11, 4049.

(61) Mueller, A. V., Eden, M. J., Oakes, J. M., Bell, C., Fernandez, L. A. Quantitative method for comparative assessment of particle filtration efficiency of fabric masks as alternatives to standard surgical masks for PPE. medRxiv. 2020, https://doi.org/10.1101/2020.04.17.20069567.

(62) Verma, S., Dhanak, M., Frankenfield, J. Visualizing the effectiveness of face masks in obstructing respiratory jets. Phys. Fluids. 2020, 32, 061708.

(63) Drewnick, F., Pikmann, J., Fachinger, F., Moormann, L., Sprang, F., Borrmann, S. Aerosol filtration efficiency of household materials for homemade face masks: influence of material properties, particle size, particle electrical charge, face velocity, and leaks. Aerosol Sci. Technol. 2020, https://doi.org/10.1080/02786826.2020.1817846.

(64) Davies, A., Thompson, K.-A., Giri, K., Kafatos, G., Walker, J., Bennett, A. Testing the efficacy of homemade masks: Would they protect in an influenza pandemic? Disaster Med. Public Health Prep. 2013, 7, 413–418.

(65) Shakya, K. M., Noyes, A., Kallin, R., Peltie, R. E. Evaluating the efficacy of cloth facemasks in reducing particulate matter exposure. J. Expo. Sci. 2017, 27, 352–357.

(66) Chu, D. K., Akl, E. A., Duda, S., Solo, K., Yaacoub, S., Schünemann H. J., on behalf of the COVID-19 Systematic Urgent Review Group Effort (SURGE) study authors. Physical distancing, face masks, and eye protection to prevent person-to-person transmission of SARS-CoV-2 and COVID-19: a systematic review and meta-analysis. Lancet 2020, 395, 1973–1987.

(67) Kai, D., Goldstein, G.-P., Morgunov, A., Nangalia, V., Rotkirch, A. Universal Masking is Urgent in the COVID-19 Pandemic: SEIR and Agent Based Models, Empirical Validation, Policy Recommendations. arXiv: 2020, 2004.13553.

(68) Watanabe, T., Bartrand, T. A., Weir, M. H., Omura, T., Haas, C. N. Development of a dose-response model for SARS Coronavirus. Risk Analysis. 2010, 30, 1129–1138.

(69) Schröder, I., COVID-19: A risk assessment perspective. Chem. Health Saf. 2020, https://dx.doi.org/10.1021/acs.chas.0c00035.

(70) Karimzadeh, S., Bhopal, R., Huy, N. T. Review of viral dynamics, exposure, infective dose, and outcome of COVID-19 caused by the SARS-COV2 virus: comparison with other respiratory viruses. Preprints 2020, 2020070613.

(71) Salomoni, S., van den Hoorn, W., Hodges, P. Breathing and Singing: Objective characterization of breathing patterns in classical singers. PLOS One. 2016, 11, e0155084.

(72) Buonanno, G., Stabile, L., Morawska, L. Estimation of airborne viral emission: Quanta emission rate of SARS-CoV-2 for infection risk assessment. Environ. Int. 2020, 141, 105794.

(73) Morawska, L., Cao, J. Airborne transmission of SARS-CoV-2: The world should face the reality. Environ. Int. 2020, 139, 105730.

(74) Fisk, W. J. The ventilation problem in schools: literature review. Indoor Air. 2017, 27, 1039–1051, https://doi.org/10.1111/ina.12403

(75) Liu, J., Ning, Z., Chen, Y., Guo, M., Liu, Y., Kumar Gali, N., Sun, L., Duan, Y., Cai, J., Westerdahl, D., et al. Aerodynamic analysis of SARS-CoV-2 in two Wuhan hospitals. Nature 2020, 557–560.

(76) Kähler, C. J., Fuchs, T., Hain, R. Können mobile Raumluftreiniger eine indirekte SARS-CoV-2 Infektionsgefahr durch Aerosole wirksam reduzieren? 2020, https://doi:10.13140/RG.2.2.27503.46243.

